# “Effectiveness of COVID-19 Vaccines in preventing Infectiousness, Hospitalization and Mortality: A Historical Cohort Study Using Iranian Registration Data During Vaccination program”

**DOI:** 10.1101/2022.01.18.22269330

**Authors:** Alireza Mirahmadizadeh, Alireza Heiran, Kamran Bagheri Lankarani, Mohammadreza Serati, Mohammad Habibi, Owrang Eilami, Fatemeh Heiran, Mohsen Moghadami

**Affiliations:** Non-communicable Diseases Research Center, Shiraz University of Medical Sciences, Shiraz, Iran; Health Policy Research Center, Shiraz University of Medical Sciences, Shiraz, Iran; Statistics and Information Technology Management, Shiraz University of Medical Sciences, Shiraz, Iran; Shiraz University of Medical Sciences, Shiraz, Iran; Medis Holding, Shiraz, Iran; University of Isfahan, Isfahan, Iran

**Keywords:** COVID-19, SARS-CoV-2, COVID 19 Vaccines, Vaccine, Effectiveness, Real-world, Cohort

## Abstract

**Background:** There are some concerns about the effectiveness of the inactivated and vector-based vaccines against SARS-CoV-2 in the real-world settings with the emergence of new mutations, especially variants of concern. Data derived from administrative repositories during mass-vaccination campaigns or programs are of interest to study vaccine effectiveness (VE).

**Methods:** Using 4-repository administrative data linkage, we conducted a historical cohort study on a target population of 3,628,857 inhabitants aged at least 18 years residing in Southern Iran.

**Results:** We estimated 71.9% [95% CI: 70.7-73.1%], 81.5% [95% CI: 79.5-83.4%], 67.5% [95% CI: 59.5-75.6%], and 86.4% [95% CI: 84.1-88.8%] hospitalization reduction for those who received the full vaccination schedule of BIBP-CorV, ChAdOx1-S/nCoV-19, rAd26-rAd5, and BIV1-CovIran vaccines, respectively. A high reduction in mortality – at least 85% – was observed in all age subgroups of fully immunized population.

**Conclusion:** The pragmatic implementation of a vaccination plan including all available vaccine options in the Iranian population was associated with a significant reduction in documented COVID-19 infection, hospitalization, and death associated with COVID-19.

**Key points:** The mass vaccination program with implementing a group of vaccines, that even for some of them (rAd26-rAd5, and BIV1-CovIran vaccines) have been only regionally authorized for emergency use, has been associated with a dramatic reduction in documented COVID-19 infection, as well as in hospitalization and deaths related to the COVID-19 diagnosis.

## 1- Introduction

Many mass-vaccination campaigns or programs are currently underway in all around the world to curb the spread of SARS-CoV-2. In the Islamic Republic of Iran, vaccination was initially started for immunocompromised patients, older people and health workers on February 9, 2021, with rAd26-rAd5 (Gam-COVID-Vac, Sputnik V). Subsequently, vaccination program was expanded with the other COVID-19 vaccines authorized by the Iranian ministry of health, including BBIBP-CorV vaccine (Sinopharm) and ChAdOx1-S/nCoV-19 vaccine (AZD1222, Oxford– AstraZeneca) ^[1-3]^ with coverage of persons in clinical risk groups and younger-age cohorts, followed by entire population above 12 years old. As of October 22, 2021, a total of 78,665,265 COVID-19 vaccine doses has been administrated nationally. Controlled clinical trials and real-world clinical studies from some countries have yielded clear evidence of the effectiveness of the just-mentioned vaccines ^[4-6]^.

However, with the emergence of new mutations, especially variants of concern (VOC), there are some debates against the protective effects of the vaccines. Laboratory findings indicate that serum samples from vaccinated persons have been attenuated for the neutralization effects against the B.1.351 (beta) variant ^[7,8]^. Moreover, observational data from Qatar showed a modestly reduced effectiveness against symptomatic infection caused by beta variant but still high levels of effectiveness against severe, critical, or fatal disease amongst people vaccinated with the BNT162b2 vaccine (Pfizer–BioNTech) ^[9]^. The delta variant is characterized by some new mutation on the spike protein ^[10]^. Some of these mutations might affect immune responses focused toward the key antigenic regions of receptor-binding protein and S1–S2 cleavage site. It appears that strains with mutations at this specific location can increase replication, leading to increased replication transmission and higher viral loads ^[11]^. Therefore, there are some concerns about the effectiveness of the available vaccine in the real-world settings with widespread distribution of delta variant. For example, researchers demonstrated an accelerated decline in protection against SARS-CoV infection by the fourth month after vaccination. Or, effectiveness has been reached low levels of approximately 20% by the seventh month after the second dose of the BNT162b2 vaccine ^[12]^.

In this study, we sought to estimate the effectiveness of the four mostly used vaccines in the Iranian national vaccination program against SARS-CoV-2, rAd26-rAd5, ChAdOx1-S/nCoV-19, BBIBP-CorV, and BIV1-CovIran (COVIran Barekat) against infection, hospitalization, and death caused by the circulating variants from February till late October in the country.

## 2- Materials and Methods

### 2-1- Study Design, Population and Data Repositories

As a potential spin-off of administrative data linkage, we conducted a historical cohort study on the individual data of a target population of 3,628,857 (out of 4,943,933) inhabitants aged at least 18 years residing in the regions under cover of the Shiraz University of Medical Sciences (SUMS), Fars province, Southern Iran, from the start of the vaccination program (February 09, 2021) to the end of follow-up (October 22, 2021). This study followed the Strengthening the Reporting of Observational Studies in Epidemiology (STROBE) reporting guideline. Biomedical Research Ethics Committees of Shiraz University of Medical Sciences gave ethical approval for this work (code = IR.SUMS.REC.1400.486) (supplementary file).

By using recoded national ID as the join variable, four data repositories were combined. All data were limited to the population under cover of SUMS, which is in charge of vaccination program, COVID-19 reverse transcription-polymerase chain reaction (RT-PCR) tests, COVID-19 hospitalization, COVID-19 outcome registry, and health data documentation.

[1] The first repository was the “Integrated Health System” (SIB, In Persian: “Samaneh Yekparche-ye Behdashti”) – an electronic health record (EHR) system established 5 years ago – that by the start of vaccination program was introduced with the vaccination data. This repository includes demographic characteristics (i.e., sex and age), address, and type, dose and data of the administered vaccine. The 4 vaccines studied against COVID-19 were [1] the BBIBP-CorV vaccine, which is a monovalent Vero cells vaccine composed of inactivated 19nCoV-CDC-Tan-HB02 (HB02) strain of SARS-CoV-2 virus antigens. ^[13]^; [2] the ChAdOx1-S/nCoV-19 vaccine, which is a modified rAd vector (ChAdOx1), containing the full-length codon-optimized coding sequence of the spike protein of SARS-CoV-2 (ChAdOx1-S/nCoV-19), with a tissue plasminogen activator leader sequence ^[14]^; [3] the rAd26-rAd5, which is a recombinant replication-deficient adenovirus (rAd)-based vaccine, containing rAd type 26 (rAd26) and rAd type 5 (rAd5) vectors that both of them carry the gene for SARS-CoV-2 full-length glycoprotein S (rAd26-S and rAd5-S) ^[2]^; and [4] the BIV1-CovIran (COVIran Barekat), which is an inactivated whole-virus SARS-CoV-2 vaccine ^[15]^. The first two vaccines have been authorized for emergency use by WHO, while the other two vaccines have been listed for emergency use, regionally, so far.

[2] The second repository was “CORONALAB” that contains the data of all people who underwent a COVID-19 RT-PCR test in public or private centers by the beginning of COVID-19 pandemic. Through this dataset, definite cases of COVID-19 infection were identified. This repository includes demographic characteristics, RT-PCR sampling date (equivalent to date of COVID-19 infection, if positive) and result, occupation (especially being a health worker), underlying medical condition (i.e., pregnancy, diabetes mellitus, malignancy, hypertension, cardiovascular disease, chronic kidney disease, and pulmonary disease), presenting sign and symptoms (i.e., nausea, diarrhea, generalized body pain, dyspnea, cough, and fever), and if a subject was hospitalized or not.

[3] The Third repository was hospitals’ “Medical Care Monitoring Center” (MCMC); that is, during COVID-19 pandemic was introduced with recording data about all suspected hospitalizations (Gray Zones admissions) due to COVID-19 as well as between-the-wards and - hospitals transfers. The acquired dataset includes admission data, underlying medical condition, presenting sign and symptoms, inpatient RT-PCR result, ward and hospital transfers’ date, outcome, and outcome data.

[4] The last repository was department of health’s “Registry of Deaths”, which served to recheck the MCMC outcome data. In addition, this was used as an adjacent dataset to add people who died within 30 days of an admission related to COVID-19 diagnosis but their data were missed in MCMC dataset. To these ends, unofficial monthly datasets were made available with a 10-day delay for research purposes, and we specifically searched for U07.1 (denoting COVID-19, virus identified) and U07.2 (denoting COVID-19, virus not identified) ICD-10 codes with date of death ^[16]^. Moreover, several other overlapping variables were rechecked using the just-mentioned repositories, including [1] demographics (using all four repositories), [2] hospitalization (using CORONALAB and MCMC), and [3] RT-PCR result (using CORONALAB and MCMC).

### 2-2- Data Processing and Outcomes

The assessed outcomes were the incidence density (count _event_/100,000 persons-day) of [1] COVID-19 infection confirmed by RT-PCR, [2] hospitalization from suspicious or confirmed COVID-19 infection lasting more than 24 hours, [3] RT-PCR-confirmed hospitalization from COVID-19, [4] death in hospital from suspicious or confirmed COVID-19 infection, [5] and RT-PCR-confirmed death in hospital from COVID-19.

Subjects who had multiple RT-PCR tests were treated in two different ways: [1] for those who had “serial” tests (e.g., subjects who required a negative result for return to works), date of the first positive test was recorded; [2] for those who had “sporadic” tests, date of all positive tests were recorded. Furthermore, hospitalization with length of stay (LoS) less than 24 hours as well as hospital readmissions within 30 days after discharge were ignored.

A subject was excluded if [1] aged <18 years at initiation of vaccination program (n = 1,315,076), [2] had previous COVID-19 infection (either positive RT-PCR test or being symptomatic), hospitalization or death prior to the initiation of vaccination program or before 14 days after receiving the second dose (n = 81,069), [3] their first follow-up day (index date) – which was 14 days after the second dose date – would be beyond the end of follow-up (October 22, 2021) (n = 955,393), [4] did not receive the second dose of vaccine (n = 585,608), [5] was not vaccinated with two doses of an identical vaccine (n = 11,226), [6] vaccinated with uncommon vaccines (n = 2,304), and [7] received the third dose of a vaccine (n = 3,908). It should be noticed that 107,201 subjects were also excluded due to unreliable data, i.e., missing or misleading vaccination date and duplicated data.

Subjects defined as vaccinated if [1] received two doses of an identical vaccine and [2] their index date was before the end of follow-up (October 22, 2021). In each vaccinated subject, persons-day was calculated by subtracting the index date from the date of censoring day, which was the occurrence date of any of the aforementioned events or, if no event was developed, the last day of follow-up (October 22, 2021).

In unvaccinated people, persons-day was calculated by subtracting index date (March 23, 2021; 42 days [assuming 4 weeks of between-doses interval + 14 days after second dose was injected] after the start of the vaccination program) from the occurrence date of any of the aforementioned events. Similar to the vaccinated group, unvaccinated people who did not develop any event, were assumed to be alive and the date of data censoring was set as October 22, 2021. Importantly, according to the Iranian notional wide vaccination program, BBIBP-CorV, rAd26-rAd5 and BIV1-CovIran vaccines required two doses separated by an interval of at least 4 weeks; but this interval was 3 months for ChAdOx1-S/nCoV-19 vaccine. Hence, for assessing the effectiveness of just-mentioned vaccine, the index date of unvaccinated cohort was set to May 24, 2021 (matched risk period).

### 2-3- Statistical Analysis

The R programing language (version 4.0.4 for MacOS) was used for statistical analysis, and its’ package “tidyverse” was utilized for data combination and data cleaning to yield an analysis-ready dataset.

To calculate vaccine effectiveness, relative risk (RR) values (with [95% confidence interval (CI)]) were subtracted from 1. We utilized the aggregated persons-day as denominator of the relative risk calculation fraction (Box 1), which is expected to correct for differences in exposure (follow-up) times, adjust for heterogenous “risk periods” (varying dynamic of a pandemic, change in dominant variants, as well as, the fact of more vaccinated people, less burden of disease), and consider the time intervals for those who developed an event, which were censored before the end of study. The confidence interval of each RR value was calculated based on the approximate procedures proposed by Ederer and Mantel ^[17]^ (Table 1).

**Table 1.** Relative risk formulas of incidence densities ^[18]^

|  |
| --- |
| 1) $RR = \frac{\frac{Event_{vaccinated}}{Aggregated\ persons - day_{vaccinated}}}{\frac{Event_{unvaccinated}}{Aggregated\ persons - day_{unvaccinated}}}$ |
| 2) $95\% CI = Lower\ limit = \left[ \frac{P_L}{1 - P_L} \right] \times \frac{L_2}{L_1}; Upper\ limit = \left[ \frac{P_U}{1 - P_U} \right] \times \frac{L_2}{L_1}$ |
| • $O_1 = events\ in\ vaccinated\ people$<br>• $O_2 = events\ in\ unvaccinated\ people$<br>• $L_1 = persons - day\ in\ vaccinated\ people$<br>• $L_2 = persons - day\ in\ unvaccinated\ people$<br>• $R_1 = event\ rate\ in\ vaccinated\ people\ (O_1/L_1)$<br>• $R_2 = event\ rate\ in\ unvaccinated\ people\ (O_2/L_2)$<br>• $\hat{P} = set\ limits\ on\ the\ ratio\ of\ events\ in\ vaccinated\ people = O_1/O_1 + O_2$<br>• $Lower\ (P_L)\ and\ Upper\ (P_U)\ limits\ of\ the\ 95\% CI = P_L = \hat{P} - \left[ 1.96 \times \sqrt{\frac{\hat{P}(1-\hat{P})}{O_1+O_2}} \right]; P_U = \hat{P} + \left[ 1.96 \times \sqrt{\frac{\hat{P}(1-\hat{P})}{O_1+O_2}} \right]$ |

Also, vaccine effectiveness was reported in different age groups (18-44, 45-64 and >64 years), separately. For this analysis, the unvaccinated cohort was selected from the same age group. Finally, the incidence density per each 100,000 persons-day of follow-up was reported. For this purpose, the “PersonTime[1] module” of “Open Source Epidemiologic Statistics for Public Health” (Open Epi) was used to calculate persons-day [95% CI] through “Mid-P exact test” with Miettinen’s modification ^[19]^.

## 3- Results

Among 1,882,148 adults (mean age: 46.74 ±17.51 years; 916,904 [48.72%] female) who composed the analysis-ready dataset, 881,638 (46.84%) were vaccinated with two doses and considered immunized, from February 09, 2021 to October 22, 2021. The mean ages were 53.39 ±15.98 and 40.80 ±16.66 years, and 369,274 (41.88%) and 547,630 (54.74%) were female in the vaccinated and unvaccinated cohorts, respectively. The vaccines composition was 75.33% for BIBP-CorV (n = 664,101), 14.87% for ChAdOx1-S/nCoV-19 (n = 131,102), 1.62% for rAd26-rAd5 (n = 14,273), and 8.18% for BIV1-CovIran (n = 72,162).

Amongst 948,230 people aged 18-44 years, 259,949 (27.41%%) received 2 doses of a vaccine. Amongst 585,166 people aged 45-64 years, 393,994 (67.33%) received 2 doses of a vaccine. And, amongst 348,752 people aged 65 years and older, 227,695 (65.29%) received 2 doses of a vaccine.

The incidence density of confirmed COVID-19 was 20.5 [95% CI: 20.3-20.7] cases/100,000 persons-day among the unvaccinated, and 4.12 [95% CI: 4.02-4.22], 3.49 [95% CI: 3.30-3.70], 5.19 [95% CI: 4.47-5.99], and 2.64 [95% CI: 2.41-2.88] cases/100,000 persons-day among those who fully immunized with BIBP-CorV, ChAdOx1-S/nCoV-19, rAd26-rAd5, and BIV1-CovIran vaccines, respectively. These yielded 79.9% [95% CI: 79.4-80.4%], 84.4% [95% CI: 83.5-85.3%], 74.7% [95% CI: 71.0-78.4%], and 87.1% [95% CI: 86.0-88.3%] positive RT-PCR tests reduction rate for those who received the full vaccination schedule of BIBP-CorV, ChAdOx1-S/nCoV-19, rAd26-rAd5, and BIV1-CovIran vaccines, respectively (Table 2).

**Table 2.** Incidence density per 100,000 persons-day and SARS-CoV-2 vaccine effectiveness regarding COVID-19 infection, hospitalization and death in the historical cohorts of fully vaccinated and nvaccinated people

| Vaccine | Infection<br>[Positive RT-PCR] | Hospitalization<br>[suspicious/definite<sup>*</sup>] | Hospitalization<br>[definite] | Death<br>[suspicious/definite] | Death<br>[definite] |
| --- | --- | --- | --- | --- | --- |
| ***Incidence density per 100,000 persons-day*** |  |  |  |  |  |
| BiBP-CorV | 4.12 [4.02-4.22]<sup>§</sup> | 2.27 [2.20-2.35] | 1.50 [1.44-1.56] | 0.17 [0.15-0.19] | 0.13 [0.12-0.15] |
| ChAdOx1-S/nCoV-19 | 3.49 [3.30-3.70] | 1.90 [1.76-2.05] | 1.10 [0.99-1.22] | 0.10 [0.07-0.13] | 0.06 [0.04-0.09] |
| rAd26-rAd5 | 5.19 [4.47-5.99] | 3.01 [2.48-3.61] | 1.74 [1.35-2.21] | 0.03 [0.00-0.13] | 0.00 [-]<sup>‡</sup> |
| BiV1-Coviran | 2.64 [2.41-2.88] | 1.20 [1.05-1.37] | 0.73 [0.61-0.86] | 0.03 [0.00-0.06] | 0.02 [0.00-0.04] |
| Unvaccinated<sup>†</sup> | 20.5 [20.3-20.7] | 8.31 [8.19-8.44] | 5.34 [5.24-5.45] | 1.81 [1.14-1.23] | 0.95 [0.91-1.00] |
| ***Effectiveness*** |  |  |  |  |  |
| BiBP-CorV | 79.9% [79.4-80.4%] | 72.6% [71.7-73.6%] | 71.9% [70.7-73.1%] | 85.3% [83.5-87.1%] | 86.1% [84.1-88.0%] |
| ChAdOx1-S/nCoV-19 | 84.4% [83.5-85.3%] | 79.4% [77.8-81.1%] | 81.5% [79.5-83.4%] | 89.5% [85.8-93.2%] | 91.8% [88.2-95.4%] |
| rAd26-rAd5 | 74.7% [71.0-78.4%] | 63.8% [57.0-70.6%] | 67.5% [59.5-75.6%] | 97.7% [93.1-100%] | 100% [-]<sup>‡</sup> |
| BiV1-Coviran | 87.1% [86.0-88.3%] | 85.5% [83.6-87.4%] | 86.4% [84.1-88.8%] | 97.7% [95.7-99.7%] | 98.3% [96.3-100%] |
| <sup>*</sup> Suspicious = Negative RT-PCR but clinically in favor of COVID-19 infection; Definite = Positive RT-PCR and clinically in favor of COVID-19 infection.<br><sup>†</sup> Incidence densities of the unvaccinated cohort for ChAdOx1-S/nCoV-19 vaccine were 22.36 [22.11-22.60], 9.25 [9.10-9.41], 5.94 [5.81-6.06], 0.92 [0.87-0.97], and 0.73 [0.69-0.78], respectively.<br><sup>§</sup> 95% confidence interval<br><sup>‡</sup> No mortality reported<br>Abbreviations: RT-PCR = reverse transcription–polymerase chain reaction. |  |  |  |  |  |

The incidence density of hospitalization with confirmed COVID-19 diagnosis was 5.34 [95% CI: 5.24-5.45] cases/100,000 persons-day among the unvaccinated, and 1.50 [95% CI: 1.44-1.56], 1.10 [95% CI: 0.99-1.22], 1.74 [95% CI: 1.35-2.21], and 0.73 [95% CI: 0.61-0.86] cases/100,000 persons-day among those who fully immunized with BIBP-CorV, ChAdOx1-S/nCoV-19, rAd26-rAd5, and BIV1-CovIran vaccines, respectively. These yielded 71.9% [95% CI: 70.7-73.1%], 81.5% [95% CI: 79.5-83.4%], 67.5% [95% CI: 59.5-75.6%], and 86.4% [95% CI: 84.1-88.8%] hospitalization reduction for those who received the full vaccination schedule of BIBP-CorV, ChAdOx1-S/nCoV-19, rAd26-rAd5, and BIV1-CovIran vaccines, respectively (Table 2).

The incidence density of death with confirmed COVID-19 diagnosis was 0.95 [95% CI: 0.91-1.00] cases/100,000 persons-day among the unvaccinated, and 0.13 [95% CI: 0.12-0.15], 0.06 [95% CI: 0.04-0.09], 0.00, and 0.02 [95% CI: 0.00-0.04] cases/100,000 persons-day among those who fully immunized with BIBP-CorV, ChAdOx1-S/nCoV-19, rAd26-rAd5, and BIV1-CovIran vaccines, respectively. These values were represented 86.1% [95% CI: 84.1-88.0%], 91.8% [95% CI: 88.2-95.4%], 100%, and 98.3% [95% CI: 96.3-100%] reduction in mortality for BIBP-CorV, ChAdOx1-S/nCoV-19, rAd26-rAd5, and BIV1-CovIran vaccines, respectively (Table 2).

In those who fully immunized with a vaccine, a high reduction in mortality (>95%) was observed in all age subgroups, except for 18-44 and >64 years age groups who received BIBP-CorV vaccine (87.3% [95% CI: 79.5-94.8%] and 86.3% [95% CI: 84.1-88.4%]). In addition, rate of reduction in hospitalization was noticeably lower in elderly (>64 years age group); that is, full vaccination schedule with BIBP-CorV, ChAdOx1-S/nCoV-19, rAd26-rAd5, and BIV1-CovIran vaccines, reduced hospitalization for 45.7% [95% CI: 42.1-49.2%], 76.0% [95% CI: 73.0-78.9%], 45.5% [95% CI: 4.9-85.8%], and 80.3% [95% CI: 72.8-87.8%], respectively. Furthermore, specifically, for the people aged >64 years, the full schedule by BIBP-CorV, ChAdOx1-S/nCoV-19, rAd26-rAd5, and BIV1-CovIran vaccines was associated with 29.1% [95% CI: 25.3-32.8%], 62.4% [95% CI: 59.0-65.6%], 76.3% [95% CI: 53.1-99.5%], and 67.0% [95% CI: 58.5-75.4%] reduction in infection (Table 3).

**Table 3.**
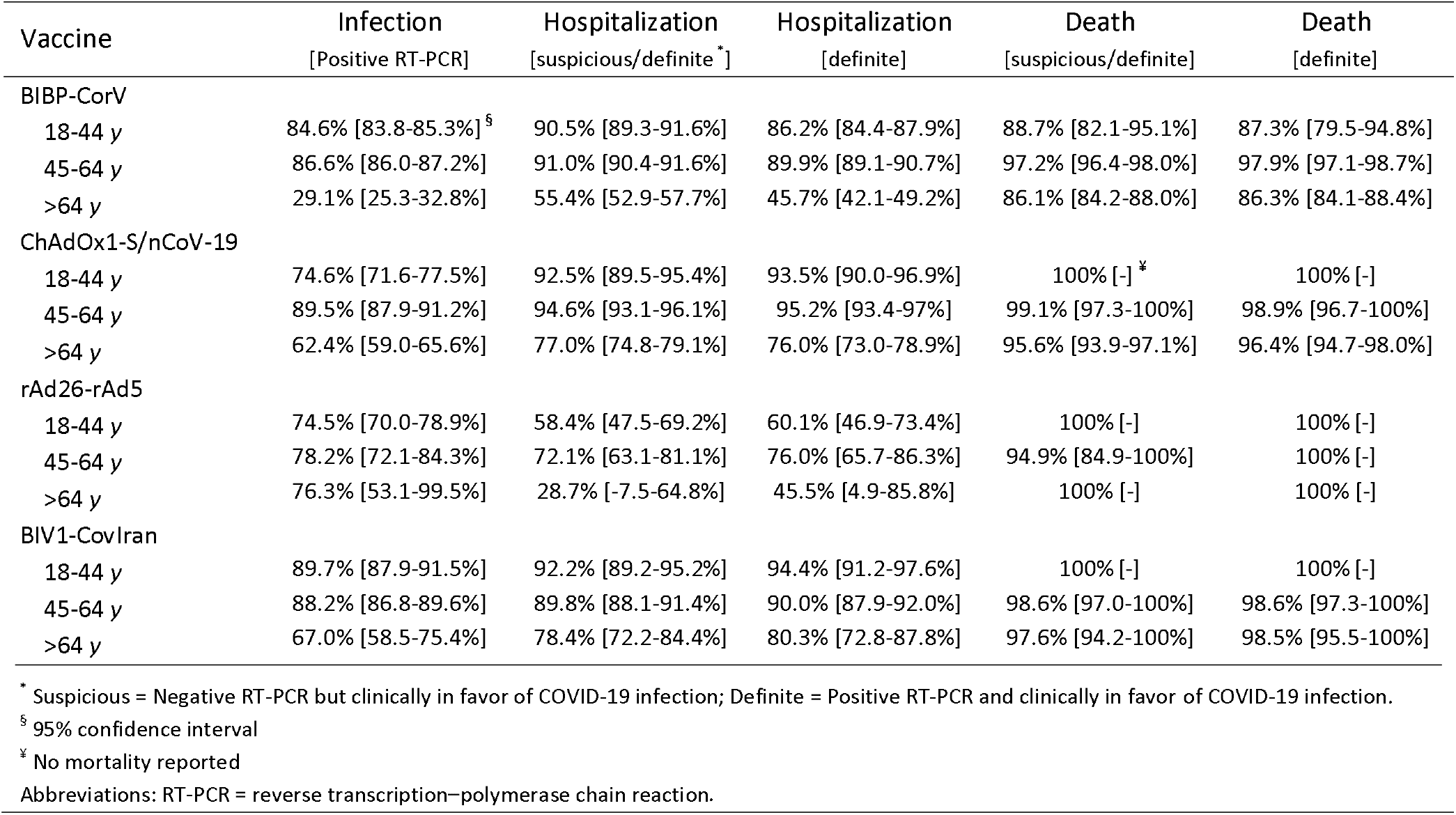
SARS-CoV-2 vaccine effectiveness in different age groups

Total and age-specific frequency of events, as well as age-specific incidence density per 100,000 persons-day, are yielded in supplementary file.

## 4- Discussion

Besides proving vaccine efficacy in clinical trials, demonstrating vaccine effectiveness (VE) in real-world settings has an essential role in strategic planning and controlling infectious diseases in the community. For various COVID-19 vaccines, there are a lot of data about efficacy ^[20-23]^. There is, to our knowledge, no VE assessment is conducted on the real-world big-data of the inactivated and vector-based vaccines that are utilizing in Islamic Republic of Iran.

In a context of vaccine shortages for our country, related to many factors (i.e., supply delays, sanctions, etc.), this study found that the mass vaccination program with implementing a group of vaccines that even for some of them (rAd26-rAd5, and BIV1-CovIran vaccines) there is limited information for effectiveness and impact, has been associated with a dramatic reduction in documented COVID-19 infection, as well as in hospitalization and deaths related to the COVID-19 diagnosis. This information is of health relevance and encourages health authorities to rapidly reach a critical mass of vaccinated population to control the disease over the country.

Our findings demonstrated high VE against laboratory-confirmed SARS-CoV-2 infection after receiving the second doses of all used vaccine. The VE point estimates reached 87.1%, with BIV1-CovIran, and at least around 75% with rAd26-rAd5. Our results consistently indicate that a high level of protection, with little variability, has been provided by all available vaccines against COVID-19 after the second dose of vaccine in the Islamic Republic of Iran. The VE estimates are consistent with the vaccine efficacy results published by the other researchers and original vaccine research groups ^[24-26]^.

In our analysis of approximately 13,987 hospitalizations of adults with laboratory-confirmed COVID-19 during February 17–October 22, 2021, receipt of 2 doses of any type of authorized vaccine was effective in preventing laboratory-confirmed COVID-19 hospitalizations among patients who were elderly (VE range = 45.5-80.3%) and those who were at younger age groups (VE range = 60.1-94.4% for 18-44-year-old age group, 76-95.2% for 45-64-year-old age group). Nonetheless, elderly was largely less protected from severe COVID-19 outcomes than younger age groups, supporting the WHO’s recommendation for administering the booster dose of a vaccine to enhance further protection, especially in elderly persons, against severe COVID-19 outcomes ^[28]^.

The analysis found significant protection associated with two doses of any of the vaccines used to prevent death from any cause. However, compared with the other used vaccines, the BBIBP-CorV vaccine was associated with a higher mortality risk during follow-up. The reasons why two doses of BBIBP-CorV were rather associated with lower protection against death than other studied vaccines remain obscure. One can explain that the low effectiveness found in people received inactivated whole virus vaccines should not be surprising, since the process of treatment used to eliminate infectivity may be effectively damaging to modify immunogenicity, especially of the antigens needed to induce cell-mediated immune responses. Additionally, it should be emphasized that although there was no large difference in the mortality among the populations that received the different types of vaccines in our study, this should be evaluated in the future studies, involving more numbers of cases – especially the other three vaccines – and other parts of the country to confirm if there is different effectiveness among these vaccines.

The findings in our study are subject to at least four limitations. First, vaccination status and outcome misclassifications might be plausible, comparing the vaccine efficacy studies, despite the high specificity of the COVID-19 vaccination status from our data repositories. Worth noting, we performed a number of data-recheck plans. Second, while our study covered a long follow-up period of 35 weeks, VE data with longer follow-up are warranted. Second, underestimation of actual SARS-CoV-2 infection among the study population might be happened due to the underuse of PCR tests for all suspected patients in the community. Third, noncomprehensive data entry – specifically, the COVID-19-related administrative data registries were not intended to collect the unvaccinated population data – for comorbidities and other confounding factors did not allow us to introduce these variables into the analysis. Fourth, due to lower numbers of vaccinated people with rAd26-rAd5 and BIV1-CovIran vaccines, VE values for these two vaccines might not be robust.

## Conclusion

The present study shows that the pragmatic implementation of a vaccination plan including all different vaccine options in the Iranian population was associated with a significant reduction in documented COVID-19 infection, hospitalization, and death associated with COVID-19. These results suggest implementing mass vaccination strategies with these available vaccines in the shortest possible time. Also, our findings are the first actual world report for an Iranian vaccine, BIV1-CovIran, with noticeable results and role in mass immunization goals.

## Supporting information

Ethics Certificate

## Data Availability

The data repositories generated during the current study are not publicly available; however, would be available from the corresponding author on reasonable request to the Shiraz University of Medical Sciences, Vice Chancellor of Research.

## Contribution

Conceptualization (A.M., A.H., K.B.L., M.M.); Data curation (M.S., M.H.); Formal analysis (A.M., O.E.); Methodology (A.H., A.M.); Project administration (A.H., F.H., M.M.); Software (A.H., F.H.); Supervision (A.M., K.B.L., M.M.); Validation (A.M., M.S., M.H.); Writing – original draft (A.H., M.M); Writing – review & editing (K.B.L., O.E.).

