## supplementary tables 1-3 for "“Effectiveness of COVID-19 Vaccines in preventing Infectiousness, Hospitalization and Mortality: A Historical Cohort Study Using Iranian Registration Data During Vaccination program”"

**Supplementary table 1.** Frequency of events regarding COVID-19 infection, hospitalization and death in the historical cohorts of fully vaccinated and unvaccinated people

| Vaccine | Infection [Positive RT-PCR] | Hospitalization [suspicious/definite *] | Hospitalization [definite] | Death [suspicious/definite] | Death [definite] |
|---|---|---|---|---|---|
| BIBP-CorV | 6,918 | 3,835 | 2,534 | 294 | 225 |
| ChAdOx1-S/nCoV-19 | 1,158 | 633 | 367 | 32 | 20 |
| rAd26-rAd5 | 181 | 109 | 63 | 1 | 0 |
| BIV1-CovIran | 482 | 221 | 133 | 5 | 3 |
| Unvaccinated † | 42,528 | 17,005 | 10,890 | 2,394 | 1,931 |

* Suspicious = Negative RT-PCR but clinically in favor of COVID-19 infection; Definite = Positive RT-PCR and clinically in favor of COVID-19 infection.
† Frequencies of the unvaccinated cohort for ChAdOx1-S/nCoV-19 vaccine were 32,702, 13,384, 8,564, 1,314, and 1,049, respectively.
Abbreviations: RT-PCR = reverse transcription–polymerase chain reaction.

**Supplementary table 2.** Age-specific frequency of events regarding COVID-19 infection, hospitalization and death in the historical cohorts of fully vaccinated and unvaccinated people

| Vaccine | Infection [Positive RT-PCR] | Hospitalization [suspicious/definite *] | Hospitalization [definite] | Death [suspicious/definite] | Death [definite] |
|---|---|---|---|---|---|
| BIBP-CorV | | | | | |
| 18-44 *y* | 1,714 | 279 | 259 | 12 | 11 |
| 45-64 *y* | 2,540 | 987 | 733 | 45 | 28 |
| >64 *y* | 2,664 | 2,089 | 1,542 | 237 | 186 |
| ChAdOx1-S/nCoV-19 | | | | | |
| 18-44 *y* | 287 | 25 | 14 | 0 | 0 |
| 45-64 *y* | 158 | 49 | 29 | 1 | 1 |
| >64 *y* | 713 | 559 | 324 | 31 | 19 |
| rAd26-rAd5 | | | | | |
| 18-44 *y* | 128 | 57 | 35 | 0 | 0 |
| 45-64 *y* | 49 | 37 | 21 | 1 | 0 |
| >64 *y* | 4 | 15 | 7 | 0 | 0 |
| BIV1-CovIran | | | | | |
| 18-44 *y* | 131 | 26 | 12 | 0 | 0 |
| 45-64 *y* | 291 | 146 | 94 | 3 | 2 |
| >64 *y* | 60 | 49 | 27 | 2 | 1 |
| Unvaccinated | | | | | |
| 18-44 *y* | 30,461 | 8,078 | 5,179 | 295 | 241 |
| 45-64 *y* | 9,190 | 5,361 | 3,541 | 797 | 658 |
| >64 *y* | 2,877 | 3,566 | 2,170 | 1,302 | 1,032 |

* Suspicious = Negative RT-PCR but clinically in favor of COVID-19 infection; Definite = Positive RT-PCR and clinically in favor of COVID-19 infection.
† Frequencies of the unvaccinated cohort for ChAdOx1-S/nCoV-19 vaccine were 23,936, 7,024, 4,551, 225, and 187, respectively.
‡ Frequencies of the unvaccinated cohort for ChAdOx1-S/nCoV-19 vaccine were 7,230, 4,401, 2,919, 524, and 436, respectively.
§ Frequencies of the unvaccinated cohort for ChAdOx1-S/nCoV-19 vaccine were 1,536, 1,959, 1,093, 565, and 426, respectively.
Abbreviations: RT-PCR = reverse transcription–polymerase chain reaction.

**Supplementary table 3.** Age-specific incidence density per 100,000 persons-day regarding COVID-19 infection, hospitalization and death in the historical cohorts of fully vaccinated and unvaccinated people

| Vaccine | Infection [Positive RT-PCR] | Hospitalization [suspicious/definite *] | Hospitalization [definite] | Death [suspicious/definite] | Death [definite] |
|---|---|---|---|---|---|
| BIBP-CorV | | | | | |
| 18-44 *y* | 3.26 [3.11-3.42] ¤ | 0.53 [0.47-0.59] | 0.49 [0.43-0.55] | 0.02 [0.12-0.39] | 0.02 [0.01-0.04] |
| 45-64 *y* | 3.09 [2.97-3.21] | 1.20 [1.12-1.27] | 0.89 [0.82-0.95] | 0.05 [0.04-0.07] | 0.03 [0.03-0.05] |
| >64 *y* | 8.02 [7.72-8.33] | 6.28 [6.01-6.55] | 4.62 [4.39-4.85] | 0.70 [0.62-0.80] | 0.55 [0.48-0.64] |
| ChAdOx1-S/nCoV-19 | | | | | |
| 18-44 *y* | 5.95 [5.29-6.67] | 0.51 [0.34-0.74] | 0.29 [0.16-0.47] | 0 [-] ¥ | 0 [-] |
| 45-64 *y* | 2.67 [2.28-3.11] | 0.83 [0.62-1.08] | 0.49 [0.33-0.69] | 0.02 [0.00-0.10] | 0.02 [0.00-0.08] |
| >64 *y* | 3.18 [2.95-3.42] | 2.49 [2.29-2.70] | 1.44 [1.29-1.60] | 0.14 [0.09-0.19] | 0.08 [0.05-0.13] |
| rAd26-rAd5 | | | | | |
| 18-44 *y* | 5.41 [4.53-6.42] | 2.30 [1.76-2.96] | 1.41 [1.00-1.94] | 0 [-] | 0 [-] |
| 45-64 *y* | 5.03 [3.76-6.59] | 3.71 [2.65-5.06] | 2.10 [1.33-3.15] | 0.10 [0.00-0.5] | 0 [-] |
| >64 *y* | 2.68 [0.85-6.46] | 10.03 [5.83-16.17] | 4.64 [2.03-9.17] | 0 [-] | 0 [-] |
| BIV1-CovIran | | | | | |
| 18-44 *y* | 2.18 [1.83-2.58] | 0.43 [0.29-0.62] | 0.20 [0.11-0.34] | 0 [-] | 0 [-] |
| 45-64 *y* | 2.72 [2.42-3.05] | 1.36 [1.15-1.60] | 0.86 [0.71-1.07] | 0.03 [0.00-0.01] | 0.02 [0.00-0.05] |
| >64 *y* | 3.73 [2.88-4.77] | 3.04 [2.28-3.99] | 1.67 [1.13-2.40] | 0.12 [0.02-0.41] | 0.06 [0.00-0.30] |
| Unvaccinated | | | | | |
| 18-44 *y* † | 21.18 [20.94-21.42] | 5.53 [5.41-5.66] | 3.54 [3.45-3.64] | 0.20 [0.18-0.23] | 0.16 [0.14-0.19] |
| 45-64 *y* ‡ | 23.06 [22.59-23.53] | 13.32 [12.96-13.68] | 8.76 [8.48-9.05] | 1.96 [1.83-2.10] | 1.62 [1.50-1.75] |
| >64 *y* § | 11.32 [10.91-11.73] | 14.07 [13.62-14.54] | 8.51 [8.16-8.87] | 5.08 [4.81-5.37] | 4.02 [3.76-4.28] |

* Suspicious = Negative RT-PCR but clinically in favor of COVID-19 infection; Definite = Positive RT-PCR and clinically in favor of COVID-19 infection.

† Incidence densities of the unvaccinated cohort for ChAdOx1-S/nCoV-19 vaccine were 23.38 [23.09-23.68], 6.79 [6.63-6.95], 4.39 [4.26-4.52], 0.22 [0.19-0.25], and 0.18 [0.15-0.21], respectively.

‡ Incidence densities of the unvaccinated cohort for ChAdOx1-S/nCoV-19 vaccine were 25.48 [24.90-26.08], 15.40 [14.95-15.86], 10.18 [9.82-10.56], 1.82 [1.67-1.98], and 1.51 [1.37-1.66], respectively.

§ Incidence densities of the unvaccinated cohort for ChAdOx1-S/nCoV-19 vaccine were 8.45 [8.04-8.89], 10.80 [10.33-11.29], 6.01 [5.66-6.37], 3.10 [2.85-3.36], and 2.33 [2.12-2.56], respectively.

¤ 95% confidence interval

¥ No mortality reported

Abbreviations: RT-PCR = reverse transcription–polymerase chain reaction.
