## Supplementary material for "“Effectiveness of COVID-19 Vaccines in preventing Infectiousness, Hospitalization and Mortality: A Historical Cohort Study Using Iranian Registration Data During Vaccination program”": Ethics Certificate

Shiraz University of Medical Sciences

### Research Ethics Committees Certificate

| Approval ID: | IR.SUMS.REC.1400.486 | Approval Date: | 2021-09-15 |
|---|---|---|---|
| Evaluated by: | Research Ethics Committees of Shiraz University of Medical Sciences | | |
| Status: | Approved | | |
| Approval Statement: | The project was found to be in accordance to the ethical principles and the national norms and standards for conducting Medical Research in Iran.<br>Notice:<br>1. Although the proposal has been approved by the Biomedical Research Ethics Committee, meeting the professional and legal requirements is the sole responsibility of the PI and other project collaborators.<br>2. This certificate is reliant on the proposal/documents received by this committee on 2021-09-15. The committee must be notified by the PI as soon as the proposal/documents are modified. | | |
| Proposal Title: | SARS-CoV-2 vaccines effectiveness against COVID-19 infection, hospitalization and mortality in a real-world setting test negative case-control study | | |
| Principal Investigator: | Name: Mohsen Moghadami<br> | | |

Dr. Mehrzad Lotfi
Committee Director
Shiraz University of Medical Sciences

Dr. Abbas Rezaianzadeh
Committee Secretary
Shiraz University of Medical Sciences
